# Prediction of fall-risk factors specific to the old-age Indian population

**DOI:** 10.1101/2023.09.05.23295087

**Authors:** Abhinav Sharma, Mehul Sharma, Ashish Arunkumar Sharma

## Abstract

**Background/Objectives:** The health care infrastructure of India, designed to treat acute problems, can benefit from preventive medicine-based policies that address chronic and non-communicable issues of relevance to India’s growing elderly population. Unintentional fall related injuries are one such issue whose economic burden can be streamlined with proper preventative public health policies. It is imperative that fall-risk factors specific to the Indian population be identified and analyzed for use in geriatric falls-risk assessment. We aim to determine factors predictive of falls in the aging Indian population in this study using Wave 1 data from the World Health Organization Study on Global Ageing and Adult Health (WHO SAGE) in India.

**Methods:** Cross-sectional analysis of results from WHO SAGE Wave 1 was conducted. Multivariate analysis was used to determine risk factors of falls specific to the Indian population in adults over the age of 50. Prediction models were created and evaluated using these risk factors and their performances were evaluated. SAGE Wave 1 India was implemented in six states that together provided nationally representative samples. Multistage stratified sampling was used to select these states and systematic sampling was used to select households from villages and urban districts within these states. Data from all individuals over the age of 50 in selected households was compiled for analysis.

**Findings:** 34 fall risk factors specific to the Indian population were determined. The model that did not weigh the factors was determined as the best model for possible use in clinically assessing old-age adults at risk for falling. Furthermore, 6 risk factors in the Indian census were used to identify falls risks hotspots on a district-level map of India.

**Conclusion:** This analysis can be used in public health policy recommendations and can form a basis for assessing and addressing falls-risk issues in India.

## Introduction

With a high crude birth rate (20.4 births per 1000 per year), low crude death rate (7.9 deaths per 1000 per year)^1^ and a 50+ age group expected to double to ∼150million^2^ by 2025, India is bound to be burdened with ailments of old age^3^. A rise in the aging population has led to an increase in non-communicable diseases (NCD) and injuries contributing to Disability-adjusted life years (DALYs); and unintentional injuries resulting from falls are now a top-25 cause of DALYs worldwide – preceding any type of cancer^4^. Falls are the 14^th^ leading cause of Years-lived with disability (YLDs) amongst the older population in India, with an estimated 800,000 YLDs in 2015^5^. Review of literature on falls in India shows that fall incidence in people aged 60+ ranged from 14-53%, and estimated that direct health care cost for a fall related hospital visit in India was ∼Rs. 45000 (∼$700)^6^.

Previous studies have associated numerous biological, environmental, behavioural, and socio- economic factors to fall risk^7,8,9^. The lack of standardized terminology, methodology and differences in sampling procedures have made cross-study comparisons challenging^10,11,12^. For example: altered gait is a well-known risk factor for falls. However, gait analysis varies between studies with some looking objectively at gait speed, while others qualitatively analyzing balance disturbances while walking. Differences in risk factors based on the geographical location/socio- economic status of a country have also been observed^8,9,11,13,14,15^. Depression, a risk factor for falls in developed countries^9^, is seldom found as a risk factor in developing countries^10^ like India^12,16,17,18,19^. These data suggest that a reliable assessment tool to weigh fall-risk factors is one that accounts for cultural, geographical and socioeconomic differences. Indeed, one recent fall assessment tool developed in Japan relied heavily on data gathered from local community institutions^13^.

We analyzed nationally representative data collected from six states in India – gathered by Wave 1 of the World Health Organization (WHO) Study on global AGEing and adult health (SAGE). Using these data, we identified non-redundant fall-risk factors (characterized in the literature or are important components of current fall-prediction tools) specific to elderly Indians. We constructed a concise risk-assessment tool that can be readily used to identify individuals at risk of falling, and used relevant data from the 2011 Indian census to visualize falls risk hotspots on a district-level map of the country.

## Methods

### Study design and Ethics Statement

This paper uses data from WHO’s SAGE. SAGE is supported by the US National Institute on Aging through Interagency Agreements OGHA 04034785; YA1323-08-CN-0020; Y1-AG-1005-0) and through research grants R01-AG034479 and R21-AG034263. All criteria in this section were fulfilled by SAGE. Of the six countries studied, we analyzed the data obtained in India. Ethics criteria was approved by the World Health Organization’s Ethics Review Board (Geneva, Switzerland), and the International Institute of Population Science’s Institutional Review Board (Mumbai, India). Retrospective analysis of the data obtained from SAGE was done after obtaining permission from SAGE India (Mumbai). The details of study design are presented in the supplementary materials.

### Patient and Public Involvement

No patient and public were involved in the design, or conduct, or reporting, or dissemination of the research.

### Detection of risk factors for fall-related injury

To better understand the aging Indian population, SAGE collected data from ∼12,000 subjects and their families using standardized household and individual questionnaires. Given our interest in studying fall-risk in the elderly, we selected for individuals over 50 years of age (n = 6263). Household and individual questions that pertained to these individuals were merged to generate a table of 1944 questions. A subject was binned as a “faller” based on the strategy described in supplementary data section: “Algorithm to determine a faller and Prevalence of Unintentional past-year fall-related injury”. Next, selection criteria described here (see also Supp. Fig. 1) was applied to clean-up redundant and low response rate questions. All questions with a response rate <20% (743 questions) were excluded. The remaining questions were manually curated, with the help of a physician, to select for variables that are important for fall-risk or meet the criteria of known risk-assessment tools (i.e. Missouri Alliance for Home Care-10, Westmead Home safety assessment). In addition, new meta-variables were generated (as described in the supplementary data section: “variable construction and selection for analysis”) to quantify cognition, waist-to-hip ratios, education etc. Odds ratios (falls vs no-falls with 95% confidence intervals) for 247 questions that met the above criteria were computed, and 65 significant questions (that had p<0.05 and were not “follow-up” questions; see supplementary data: “Variable construction and selection for analysis”) were selected.

### Risk Assessment Tool for fall-related injury

In order to build concise criteria for assessing fall-risk in the field, the following steps were taken (Supp. Fig. 1): the cohort was randomly sampled into “training” (80% of data) and “testing” (20% of data) sets.

Training set: Responses to all 65 questions above were converted into a binary output using the best Youden’s index cut-off (see Supplementary section: “Youdens cut-off for Risk Assessment”). Features that best predicted falls (binomially distributed) in the training set were determined using Least Absolute Shrinkage and Selection Operator (LASSO), where features within 1 standard error of the highest AUC (Area under the curve) were selected after a k-fold cross-validation (k = 100; implemented using glmnet package). The 34 selected features were weighted using four model construction algorithms (i.e. unweighted, logistic regression coefficients, variable importance and beta coefficient) described in the supplementary section: “construction and evaluation of fall prediction models”. The ROC curve of this data was generated and the “best” specificity, sensitivity and threshold scores for each model were determined using the pROC package.

Testing set: Youden’s index cut-off values determined on the training set were applied to their respective features, and each of the 4 models were used to predict falls. The sensitivity and specificity (i.e. the credibility) were assessed and reported at the “best” training set threshold value. Data and statistical analysis, model building and assessment of models was done using R (https://www.r-project.org).

### Geographical prevalence of fall-risk factors in India

Data on the following determined fall-risk factors are presented in the 2011 Indian census: hearing difficulty, vision difficulty, education level below primary school, gender, marital status and not having worked in the last 12 months. The prevalence of these variables was calculated in people over the age of 50 in each of the 640 districts of India. Multiple logistic regression followed by dominance analysis on these 6 variables was done in the WHO-SAGE dataset^20^, to determine relative importance weights for each variable. Relative importance weights were applied to prevalence rates for each factor to get weighted prevalence rates, that were summed for each district to get final risk-scores.

Previously, Geographic Information System (GIS) software has been used to display district-level disability prevalence using the Indian census data^21^. We used an online GIS district level map, obtained from S. Anand with permission, and modified it to display the distribution of the final risk-scores across India (https://upload.wikimedia.org/wikipedia/commons/8/89/Official_India_Map_with_Districts_201 1_Census.svg). The six states surveyed by WHO were traced on the map of India. ‘Prevalence’ and ‘Relative Risk’ are displayed for these states. Prevalence: The falls-injury prevalence rate in that state was calculated from the WHO-SAGE data set. Relative Risk: Average relative risk for these six states was calculated by obtaining the mean risk score (as described above) of all the districts in the state.

## Results

### Proportion of fall-related injuries steadily increases after 50 years of age

Unintentional fall related injuries ranged from 1-9% in various age groups in India (Figure 0). The above 80 age group displayed the highest rate of falls at 8.84%. Whereas, less than 20-year olds displayed the lowest rate of falls at 1.13%. Despite a lack of significant differences between age groups, the rate of fall related-injury steadily rose after the age of 50. The rate of injury was 4.3% in the 50-60 age group, 4.7% in the 60-70 age group and 6.5% in the 70-80 age group. We observed a jump in the fall injuries in 3 specific age groups: the 20-30 age group (p=0.08), the 50-60 age group (p=0.15), the 70-80 age group (p=0.07). Therefore, we chose age 50 as the cut- off to study factors associated with unintentional fall related injuries in the Indian population.

### Motility, pain in joints and lung dysfunction is predictive of falls in the Indian elderly

Using the strategy described above (see methods), the ∼2000 question WHO survey was shortened to 274 non-redundant, clinically/epidemiologically relevant questions with high response rate. A univariate odds ratio analysis was used to obtain 65 questions that were significantly associated with falls (p<0.05) (Fig. 1a). Although mental health (e.g. sadness), physical characteristics (e.g. waist-to-hip ratio, gender) and socioeconomic status (e.g. marriage, debt) were implicated; a vast proportion of questions associated with falls could broadly be characterized under the umbrella of “physical health” (Fig. 1b - left) and included questions that assessed overall fitness (i.e. daily exercise), joint pain/aches/arthritis and breathing difficulties (Fig. 1b - right). Overall, these data suggest that a diverse set of issues (led by joint and lung dysfunction) play a role in driving fall incidence in the Indian elderly. In order to rectify these issues, we sought to identify the individuals most at-risk of falling using a concise and accurate questionnaire that could be quickly used as a tool in the field.

**Figure 1:**
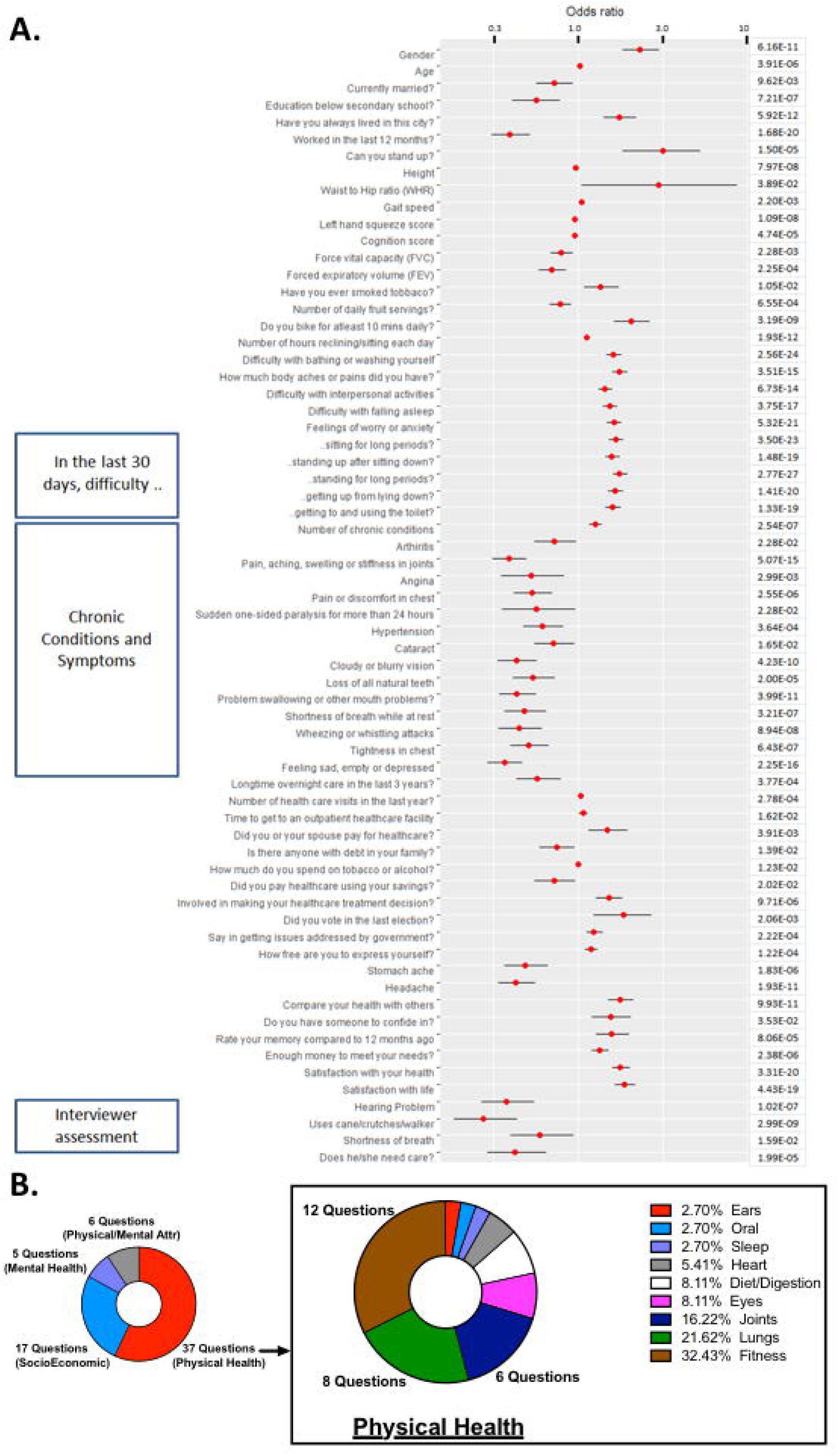
Univariate non-redundant fall-risk predictors in the elderly Indian population. A) Odds ratio and p-values for 65 significant questions (p<0.05) of the 274 non-redundant questions (see methods) from the Individual and Household surveys of the WHO-SAGE study. B) (left) Manually annotated primary categories that define the 65 univariate questions in figure 1A, and (right) distribution of physical health questions into secondary subcategories highlight the role of lung and joint dysfunction in predicting fall-risk.

### A diverse yet readily-applicable questionnaire predicts falls with an accuracy of ∼80%

To generate a short questionnaire that could easily predict falls, we applied the strategy described in the “Risk Assessment Tool for fall-related injury” section (see methods) to the 65 question highlighted above. Briefly, the data was divided into a training (80%) and a test (20%) cohort and a binary response matrix of the 65 questions was generated on the training set. A well-characterized feature selection approach (i.e. LASSO) was applied to this set, and a 34 feature-set (with the highest AUC) that best predicted falls was identified (Fig. 2a). These questions predominantly assessed overall fitness (i.e. lack of physical activity), mental health (e.g. sadness, worry) and social well-being (e.g. participation in government). Next, four weighted models (unweighted, logistic, variable importance and b-coefficient) were applied to the feature-set (weighing criteria described in supplementary data). Each of these models was bootstrapped and a confidence interval value was obtained (Supp. Table 2 and 3). Upon validating the models on the test cohort (Youden’s binary cutoff was obtained from the training set; see methods), we observed that the unweighted model performed better than all models (Fig. 2b) with an AUC of 0.81 and sensitivity of 86%. Features used to construct this model (Fig. 2c) were dominated by attributes of physical health (i.e. overall fitness and joint pain). However, mental/social health (i.e. general sadness, lack of community involvement) and personal characteristics (i.e. gender and waist-to-hip ratio) also proved to be important. The set of questions listed here (Fig. 2c), can be used to accurately predict fall-risk and prescribe future interventions.

**Figure 2:**
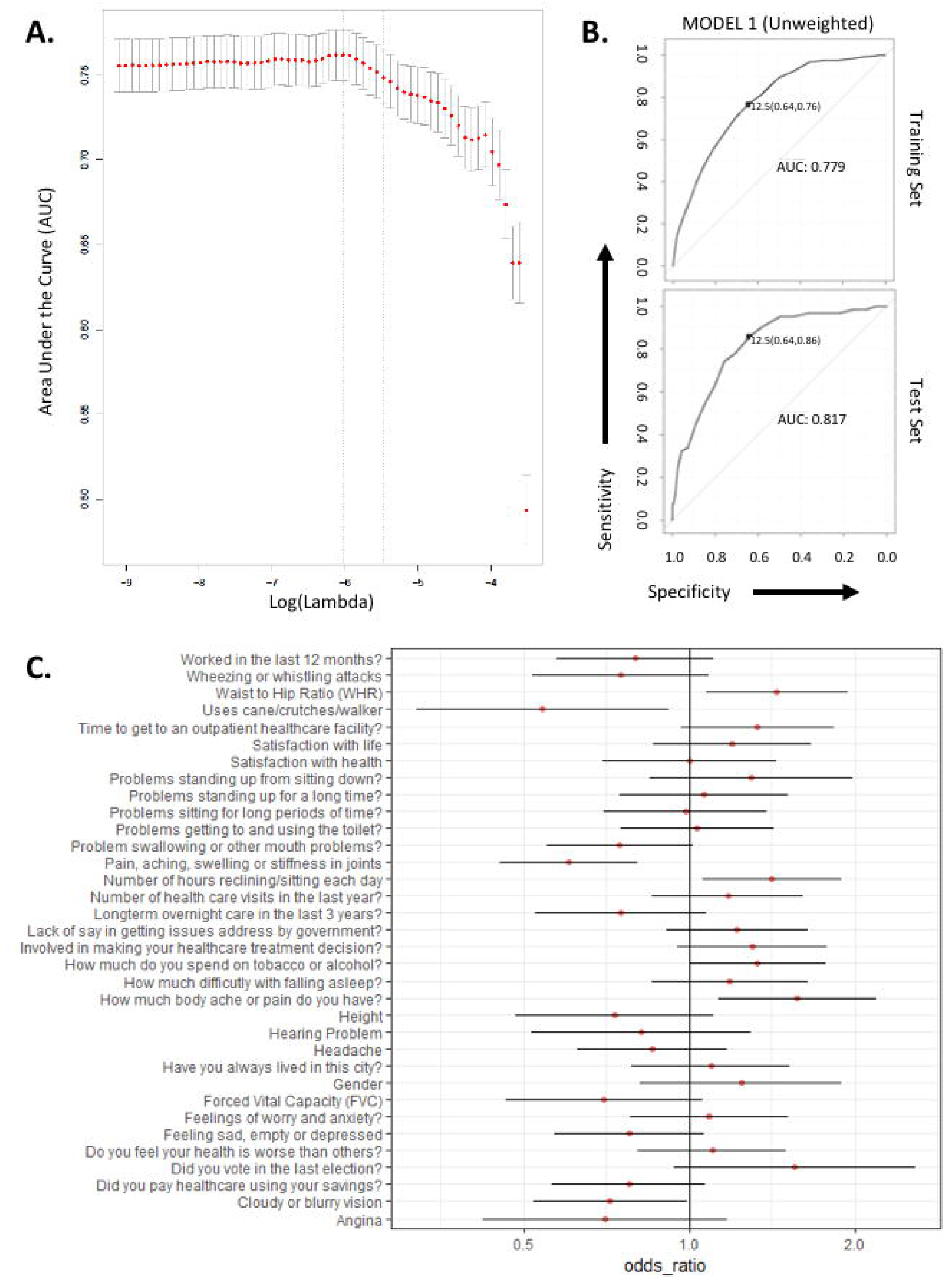
Concise and field-ready questionnaire to assess fall-risk in the elderly Indian population. A) Feature selection was done to pick the best predictors of fall-risk from the parameters listed in Figure 1A, using LASSO on binomial response and predictor variables. The AUC values for each lambda post-k-fold validation (k = 100) is shown; lambda at max AUC and 1 standard error margins are shown using vertical lines. B) Receiver Operating characteristic (ROC) curves of unweighted (Model 1 – best of 4 models) model on the “training” (top) and the “test” set are shown - the best AUC, sensitivity and specificity of the “training” set is shown. The threshold for the best training set AUC is then applied to the “test” ROC and respective sensitivity, specificity and AUC of the “test” data are also shown (bottom). C) Odds ratio and P- values are listed for 34 variables chosen using the feature selection approach in Figure 2A.

### Identification of fall-risk regions in India, using the Indian census data

Relative risk for people aged 50+ (Figure 3) shows the vulnerability of each district to falls- injury compared to the rest of India. Prevalence of fall-injury from WHO-SAGE data set showed West Bengal with the highest prevalence at 9.30%. In line with the high prevalence, the risk value for West Bengal (0.0161) was the highest compared to the six states studied. Karnataka also had a high prevalence rate of fall-injury, 5.67%, along with having a high-risk value of 0.0067. Assam and Rajasthan had lower falls-injury prevalence, 2.81% and 3.57% respectively, and also low risk values, -0.0061 and -0.0044 respectively. Although our risk values were usually in line with the census data, Uttar Pradesh did prove to be an anomaly with a high prevalence rate (6.35%), and a risk val (-0.0011) on the lower end of the spectrum (Figure 3). We believe that addition of more variables (highlighted in the assessment tool) could increase our ability to more accurately determine fall-prevalence. A possible future public health intervention which aims to acquire these additional variables via changes in the Indian census might be worth considering.

**Figure 3:**
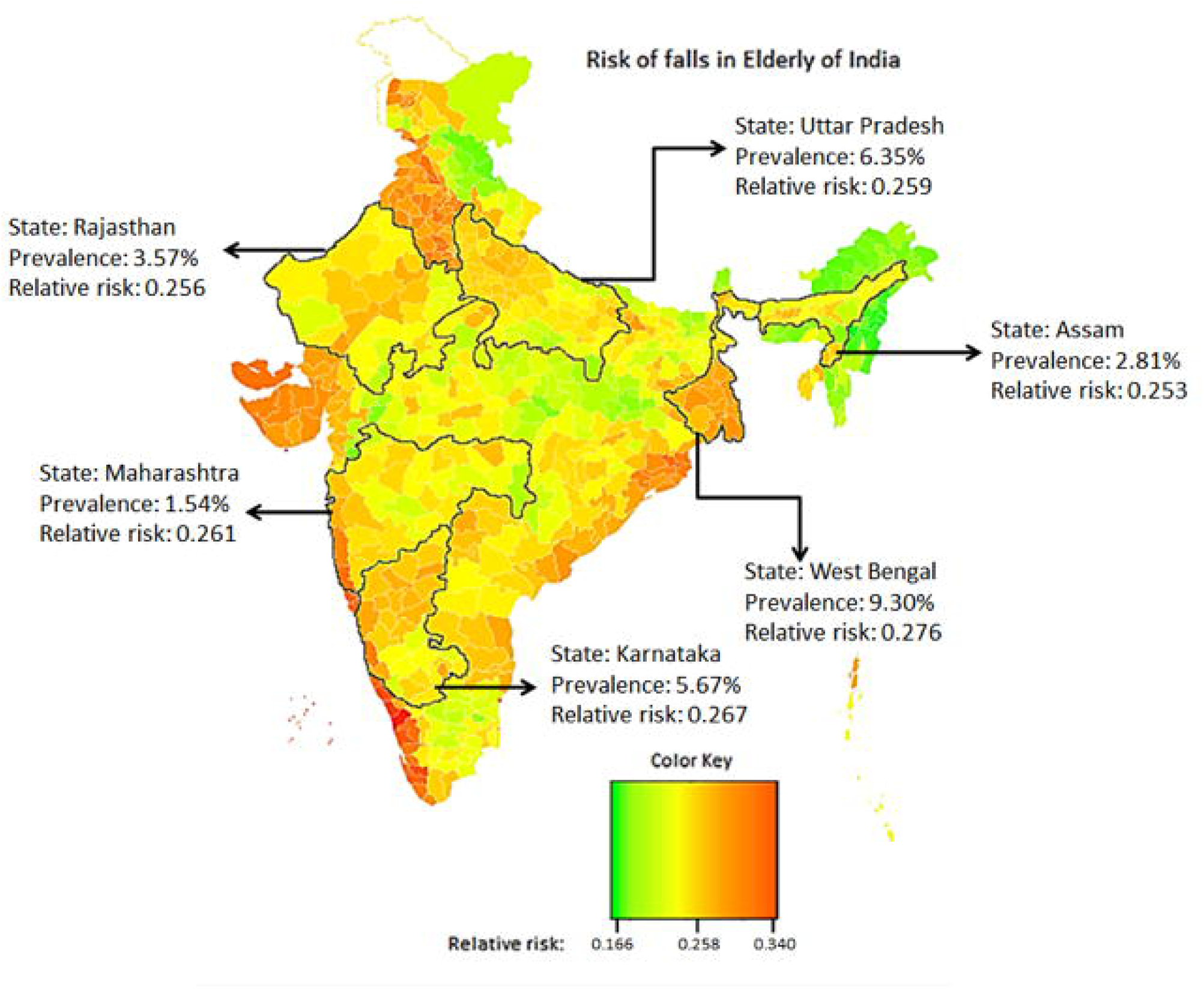
Heat map of relative risk of falls among adults age 50 or older in the districts of India. Relative importance of 6 variables (obtained from the consensus) was calculated (from SAGE- WAVE1 dataset) and applied to the prevalence of these variables in each district (as tabulated from the Indian census data). This weighted prevalence was added to obtain “relative risk” for each district. In addition, the six states studied in the SAGE-Wave1 are appropriately labelled with the prevalence of falls-injury as reported in the dataset.

## Discussion

### Strengths and weaknesses of the study

This study was a cross-sectional analysis of WHO SAGE-Wave 1 surveys conducted in India between the years 2007 – 2010. These were well-designed surveys with standardized questions that obtained data regarding most old age issues, and were carefully planned to gather nationally representative data from 6 states of India. This study analyzes the responses of 6263 individuals over the age of 50 who were part of WHOs SAGE-Wave 1. This is the first study to have analyzed the issue of falls with a sample size as large as this in India, and the first study that has analyzed the risk factors for falls collectively looking at the whole Indian population. Identifying new risk factors for falls and confirming known risk factors are important for having an informed old age population in India. This study further evaluates the accuracy of sophisticated machine learning models that use identified risk factors for the purpose of identifying an individual at a risk for falling. As per our knowledge, this is the first study to apply this methodological approach in the context of fall prevention, and exploits the standardized and generalizable nature of the survey questions by using them as variables. This study is also the first to incorporate data from the Indian census to obtain a broader picture of geographical distribution of old age issues in India, using a few identified risk factors for falls.

Although the study benefits from an extensive data set and sophisticated tools, the models created need to be validated on an external data set for further confirmation of our analysis. This can be readily tested on SAGE-Wave 2 in India, when the data becomes available publicly. The cross-sectional design of the study also restricts the drawing of any causal relationships for fall risk factors. Finally, since the study wasn’t explicitly designed to study falls, some well published risk factors for falls were not assessed. Clinical information on prior history of falls (recurrent or first time), postural hypotension, medication use and environment factors (like footwear, lighting etc.) – identified in MAHC-10 and Westmead Home Safety Assessment guide^22,23^ – are used to assess fall-risk in developed countries and might have been improved our models performance. Accurately evaluating geriatric depression^24^, cognition^25,26^ and fear of falls^27^ could have further improved our overall risk assessment. This information might have benefited the models.

### Strengths and weaknesses in relation to other studies, discussing important differences in results

Incidence of falls and risk factors associated with falls vary between countries^10^. The high number of old age homes, retirement communities and proper primary and auxiliary healthcare facilities in developed countries help in maintaining healthy living standards past 65 years of age. Consequently, a cut-off of 65 years of age is used to define and study falls (and other health issues) in the elderly. In contrast, the lack of proper healthcare facilities and unfavourable socio- economic conditions can drive quicker aging in developing and underdeveloped countries.

Indeed, telomere length, a proxy for biological ageing, has been shown to be lower in harsh socioeconomic conditions^28–30^. These data propelled us to make a case for studying fall-risk in an Indian population that is biologically younger and yet more prone to falls. Given that the average Indian life expectancy at the time of this survey was 65 years of age^31^, we decided to study fall related injuries in the aging Indian population at and after the second “jump in fall injuries’’ mark (50-60 years, Figure 0). WHO’s SAGE team has also studied ageing by gathering data from people aged 50+ in six LMIC (Low-Middle income countries), and a 50 years of age cut-off has also been used in studying fall-risks in other developing countries^11^.

We observed that incidence rates of fall-related injury varied between the six states, from 9.3% in West Bengal to 1.5% in Maharashtra (Figure 3). There already exists a sparsity of data on fall-related studies in developing countries, and the few that analyze the Indian population mostly focus on regions in northern parts of India (states of Punjab and Haryana) or southern parts (states like Kerala and Karnataka)^6^, but fail to cover several densely populated poorer states (like Uttar Pradesh, West Bengal and Assam). By contrast, our analysis is nationally representative. Furthermore, our analysis identifies possible districts in India that might have a high fall risk and those with a very low fall risk (Figure 3). Future studies looking at old-age population dynamics in geographically diverse locations can provide insight into local cultural practices that are protective against fall-related injuries.

Commonly identified risk factors for falls in developed countries like gender^32^, age^33^, education^34^, self-perceived health status^8,9^, use of assistive devices^9^, chronic conditions^8,35^, general pain^9^, lack of proper sleep, vision impairment^36^, hearing impairment^36^, gait^36^, cognition^36^, depression^37^, and ability/energy for day to day activities^9^ were all identified as potential risk factors for unintentional falls in our analyses as well. Specifically, several markers of deteriorating personal health and lack of activity contributed to fall-risk (Figure 1). However, external factors like ownership of goods and assets, dwelling characteristics and other factors generally associated with falls in LMIC (low and middle-income countries)^11^, were not associated with fall-risk. Given that physiological risk factors are more pronounced amongst recurrent fallers in India^19^, it is possible that many of the people surveyed in our study sustain injuries following recurrent falls. Living alone, another commonly identified fall-risk factor^6,7^, was not significantly correlated with fall related-injury in India (p=0.805). This could be because very few old age Indians live alone, 5%^38^, compared to 28% of American seniors^39^.

Clinically, arthritis and hypertension were identified as risk factors for falls in univariate analysis. More importantly, waist to hip ratio (WHR), was identified as a risk factor that retained its significance even after multivariate regression (Figure 2). We also found that all symptoms indicative of asthma, chronic obstructive pulmonary disease (except percent of Forced Expiratory Volume, p=0.176) and stroke incidence were strongly correlated with fall-risk, although self-reported diagnosis of these conditions was not significant. This is puzzling and could be indicative of lack of personal health education in the population. Further research is warranted to better understand accessibility to diagnostic care in India. On top of the risk factors identified above: hearing, vision, nutrition intake (fruit servings), money spent on alcohol abuse and oral health issues were also associated with fall-risk (Figure 1) and are generally known to be associated with aging.

### Possible explanations and implications for clinicians and policymakers; and unanswered questions for future research

Risk factors specific to the Indian population identified in this study will allow for the development of tailored intervention strategies. We believe that these interventions should be conducted on an individual level and will range from clinical interventions for hearing, vision and dental problems, to education and guidance for nutrition and alcohol use. Strength and balance training, which integrate traditional exercises like yoga^40,41^, might be important interventions for building strength. Multifaceted interventions, targeting individual risk factors, have previously been shown to be effective^42,43^. Although there is some uncertainty associated with the effectiveness of these interventions as compared to exercise-only interventions^44^, they address many geriatric problems contributing to an overall healthy ageing process^45^.

Our analyses reveal that factors that predict falls are primarily descriptors of physical health – 57% of the fall-risk identifying questions (Figure 1). Interestingly, of the 65 univariate variables identified the 34 that best predicted falls largely pertained to overall fitness (e.g. lethargy) and joint/body aches, possibly arising from natural/manmade infrastructural differences, in a young society (average age in India: <35 years) that does not accommodate to the elderly population. Although the fall-risk factors identified are specific to the Indian population, the methodology used to identify these variables and construct predictive models can have implications beyond India. Our approach can identify risk factors in geographical locations with specific cultural practices and limited accessibility to resources, thus improving the scope of precision public health. Importantly, the predictive models generated can be used as a quick diagnostic tool in field setting without the need of highly trained medical professionals. Even if no subsequent fall prevention public health interventions are implemented, this readily available and affordable tool can make one aware of their fall risk; which is key when addressing population health issues. We believe that diagnostic tools like the one described are critical for future interventions aimed at implementing fall prevention strategies in developing countries.

## Conclusion statement

Results from this study highlight the core risk factors associated with falls in the old age (defined as anyone over the age of 50) population of India. The study also creates a model to identify an individual at risk of falling, which can be implemented as a cheap, effective clinical tool on the field in India.

## Supporting information

All supplementary figures and tables for the manuscript

## Data Availability

All data produced in the present study are available upon reasonable request to the authors

## Author contribution

All authors contributed equally to the data analysis and manuscript preparation for this study. Dr. Abhinav Sharma is a medical doctor who was involved in manually picking the clinically relevant risk factors. Dr. Ashish Sharma and Mr. Mehul Sharma were involved in study design.

