## Supplementary material for "Prediction of fall-risk factors specific to the old-age Indian population": All supplementary figures and tables for the manuscript

**Supplementary Data**

**Methods:**

*Study Design*

SAGE Wave 1 India was implemented in six states that together provided nationally representative samples. Multistage stratified sampling was used to select these states. First, only states with populations greater than 5 million were considered for selection. Next, six specific states were selected based on level of development as determined by economic and social indicators, and geographic location within the country. One state was selected from each geographical region as well as each level of development. The list of states that met the above cut-off included: Assam, Karnataka, Maharashtra, Rajasthan, Uttar Pradesh and West Bengal. The states were further stratified based on the type of locality (rural or urban). Probability proportional to size (PPS) method was used in each state to select the primary sampling units (PSU; villages in rural locations) and census enumeration blocks (CEB; in urban locations). Systemic sampling was used to select 28 households from each village and 33 from each CEB. The numbers of rural and urban households were proportional to the population of rural and urban centres in that state.

All data was collected via standardized household and individual questionnaires. All individuals over the age of 50 in selected households were invited to complete the individual questionnaire; in parallel each participating household was invited to complete a household questionnaire. The questionnaires were translated and validated in the local language and further validated after back translation. Overall, data from 11798 individuals was obtained.

*Algorithm to determine a "faller" and Prevalence of Unintentional past-year fall-related injury*

To determine prevalence of past-year fall related injuries in different age groups, the variable 'fall-related injury' was derived from responses to questions from the SAGE Wave 1 questionnaire as described (Stewart Williams et al. 2015).

Individuals were asked "In the last 12 months, have you had any other event (other than a road traffic accident) where you suffered from bodily injury?" If the response was 'yes' then they were asked: "What was the cause of this injury?" If "fall" was selected from a list of possible responses (e.g.: struck by a person or object, stabbed, gun shot, animal bite, electric shock or fall) then the individuals were asked to confirm whether this was an 'unintentional' injury. Thereby the variable 'unintentional past-year fall-related injury' was specified.

Two cohorts of the populations were created, those that suffered fall-related injuries and those that didn't. The cohort that suffered unintentional fall injuries was compared to the rest of the population in 10-year age categories. A chi-squared test was then used to check for significant differences between age-groups. We observed an increase in falls after 50 years of age.

Subsequently, participants over 50 years of age were marked to represent the aging population of India and were analyzed in this study. In line with our study, the authors of SAGE studied aging in developing countries (where living conditions and life-expectancy are hampered by numerous socio-economic factors), specifically in people over 50 years of age.

*Variable construction and selection for analysis*

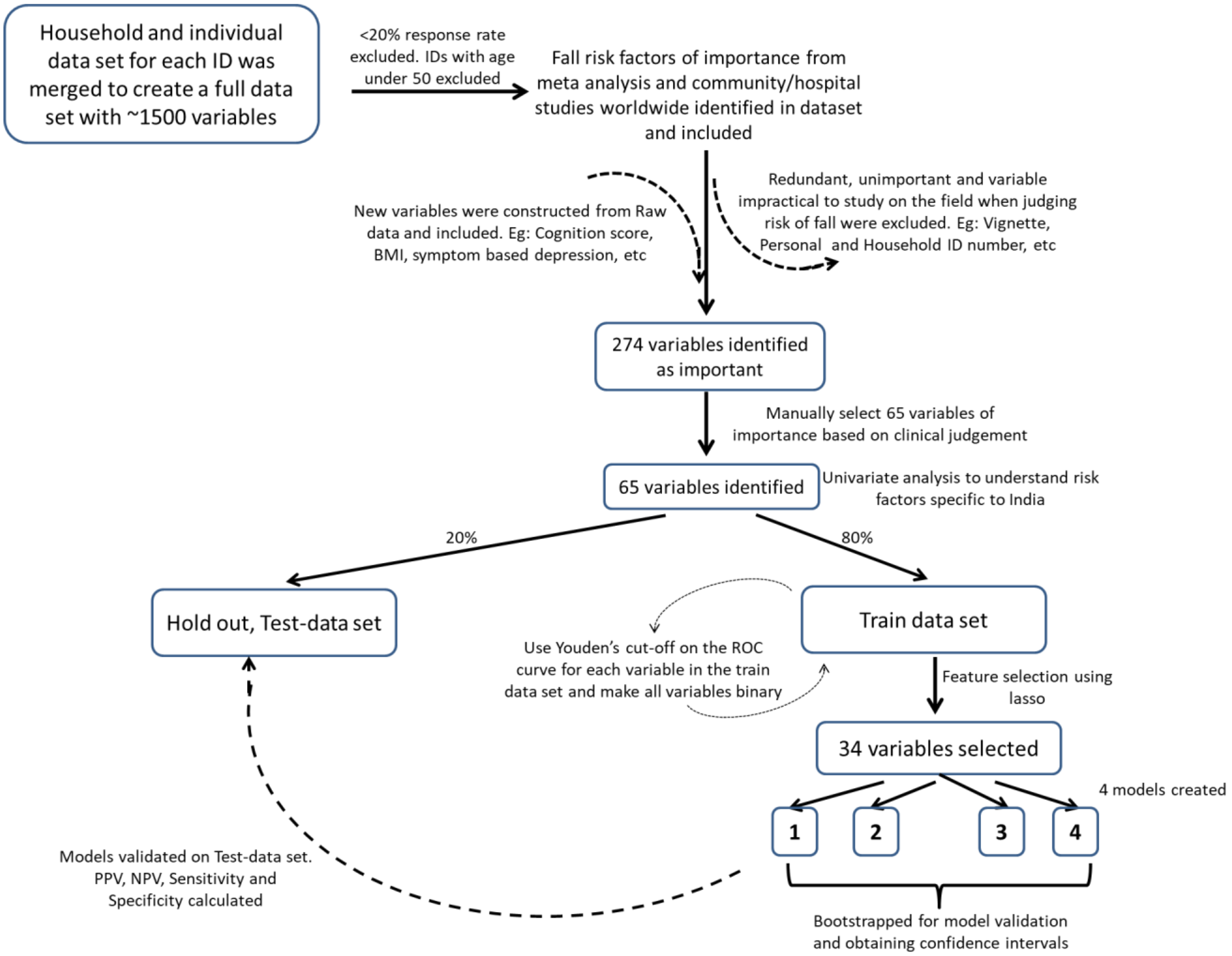

**Supplementary Figure 1. Strategy schematic for variable selection, model construction and validation**

Using individual and household identifiers provided in the WHO questionnaire, individual and household datasets were merged. The data was further cleaned by removing questions that had a response rate of less than 20%. Some variables were altered to better explain the raw data in the questionnaire.

Cognition variable was computed as described (Stewart Williams et al. 2015). Briefly, scores of verbal recall, digit span (forward and backwards) and verbal fluency questions were summed to get a cognition score. Weight (kg) and height (m) was used to calculate BMI (Himes and Reynolds 2012). Waist to hip ratio (WHR), Forced Expiratory Volume 1 (FEV1%), mean grip strength for each hand and number of self-reported diagnosed chronic conditions were calculated. Variables for symptom based diagnosis of arthritis and depression were constructed as described (Moussavi et al. 2007). Number of people in the household, and households with single inhabitant was tabulated. Highest education level in household, household family member composition (son/daughter present or grandchild present), access to water source (inside vs outside the house) and type of cooking fuel (gas, electric or kerosene stove vs others) were also calculated from the raw data. New variables for education status, marital status and Work status were constructed to match responses seen in 2011 Indian census. Education level was analyzed by categorizing the population as having more than primary education or primary education and below. Marital status was computed so that responses fell into either "currently married" or "currently unmarried". Timeline of work history was used to determine responses for the variable "worked in the last 12 months".

If follow-up to a question was found significant but the initial inquiry was not, the question was not applicable. For example: "Has anyone in the household received any financial or in-kind support from your family (children, siblings or parents) and relatives (other kin) who do not live with you?" was not found to be significant. However, the follow-up question, "What type of financial or in-kind support did your household receive in the value of food or other goods (that is, non-monetary)?" was significant but not considered.

*Construction and evaluation of fall prediction models*

1) No weights were assigned to variables and final score for an individual was computed out of 34 by summing the response to each feature/variable. 2) The estimated coefficients from multiple logistic regression were used as weights (implemented using glment package in R). 3) Variable importance was determined as the absolute z-statistic for the Wald-test, normalized to get a maximum score of 100 (implemented using varImp function in the caret package in R). 4) Standardized Beta Coefficients were used to determine variable weights (implemented using beta function in the reghelper package in R). Sensitivity, specificity, Positive Predictive Value (PPV%) and Negative Predictive Value (NPV%) were calculated as the cut-off score in the test data set. This was iterated 1000 times for each model, and the mean values were presented.

Data and statistical analysis, model building and assessment of models was done using R.

*Youdens Cutoff for Risk Assessment Tool for fall-related injury*

Each variable had a cut-off value set based on the Youden index from the Receiver Operating Characteristic (ROC) curve of that variable. If an individual scored above the cut-off score for a question then they were considered at risk for that variable. For questions with "Yes/No" responses: a negative response (one that makes an individual liable to fall) was scored as an individual at risk.

**Results:**

<u>*Socio demographic characteristics of participants that suffered fall related injuries*</u>

**Supplementary Table I: Socio Demographic differences of subjects that suffered fall related injuries and were 50+ years of age.**

| | All participants | | Age 50+ participants | |
|---|---|---|---|---|
| | *Fallers* | *Non-Fallers* | *Fallers* | *Non-Fallers* |
| Number of Observations | 639 | 11559 | 294 | 5969 |
| Average Age | 53.04 | 49.78 | 64.14 | 61.75 |
| | *% Fallers* | *% Non-Fallers* | *% Fallers* | *% Non-Fallers* |
| Male | 31.77 | 38.95 | 30.95 | 51.00 |
| Female | 68.23 | 61.05 | 69.05 | 49.00 |
| Currently Working | 37.25 | 38.97 | 22.11 | 40.86 |
| Not Working currently, but have worked in the past | 25.51 | 21.47 | 34.35 | 31.30 |
| Never Worked | 37.25 | 39.56 | 43.54 | 27.84 |
| Rural | 79.97 | 73.98 | 78.23 | 73.95 |
| Urban | 20.03 | 26.02 | 21.77 | 26.05 |
| Currently Married | 74.33 | 77.77 | 67.69 | 74.48 |
| Never married/ Separated/ Divorced/ Widowed | 25.67 | 22.23 | 32.31 | 25.52 |
| No School | 49.45 | 49.61 | 57.14 | 51.11 |
| Less than primary | 13.15 | 9.44 | 13.95 | 11.16 |
| Primary school completed | 15.96 | 13.95 | 13.95 | 14.01 |
| Secondary school completed | 9.39 | 11.55 | 6.12 | 10.14 |
| High school completed | 7.67 | 9.94 | 4.76 | 8.53 |

| | | | | |
|---|---|---|---|---|
| College completed | 3.13 | 4.07 | 3.06 | 3.48 |
| Post-grad degree | 1.25 | 1.44 | 1.02 | 1.57 |

All participants were separated between those that suffered unintentional fall related injuries and those that didn't. There were a higher percentage of women in the falls group (68.23%) vs the non-falls group (61.05%). Percent of people currently working was similar between the two groups, 37.25% in the falls group and 38.97% in the non-falls group. Education levels were also similar. 49.45% in the falls group and 49.61% of participants in the non-falls group had no schooling. The category of education with the greatest representation was primary education in both groups, 15.96% in the falls group and 13.95% in the non-falls group.

Over the age of 50, the average age of the fallers was 64.3, compared to 61.8 for non-fallers. There were a higher proportion of females in the fallers group (69.05%) compared to males (30.95%). 57.14% of adults in the falls group had no schooling and 13.95% had less than primary education, whereas in the non-falls group 51.11% of adults had no schooling and 11.16% had less than primary education. 22.11% of fallers were currently working as compared to 40.86% of non-fallers. The percentage of fallers who dwell in rural locations vs urban location was similar in the overall demographic as well as in the ages 50+ demographic: 79.97% of fallers vs 73.98% of non-fallers were rural residents for the overall demographic, and 78.23% of fallers vs 73.95% of non-fallers were rural residents in ages 50+ demographic. Although these socio-demographic factors broadly pointed out the general correlates associated with falls, they are unable to highlight the population that is more likely to fall. Next, we used the data obtained from the WHO questionnaire to understand the complex interplay of attributes that make an individual more likely to fall.

*Proportion of fall-related injuries steadily increases after 50 years of age*

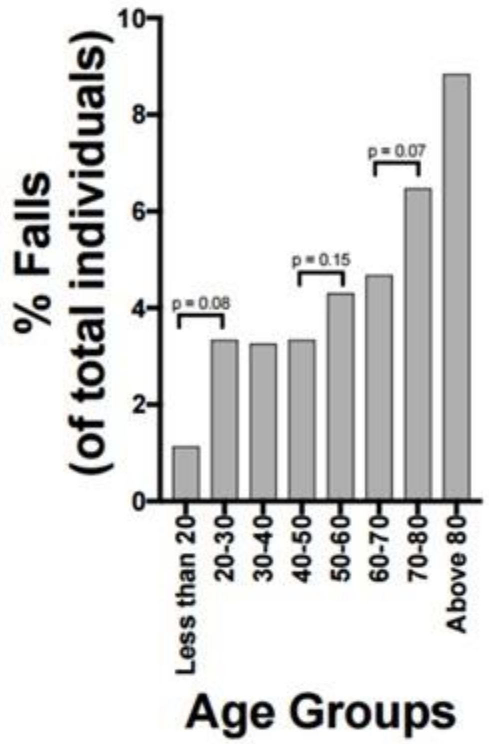

**Supplementary Figure 2: Prevalence of unintentional fall-related in India. The percentage of people that suffered accidental fall related injuries were calculated as described in the methods. The resulting data, distributed amongst 10-yr age categories, showed a jump in fall related injuries after 50 years of age, and further after 70 years of age.**

Incidence of falls and risk factors associated with falls vary between countries (Kalula et al. 2011). The high number of old age homes, retirement communities and proper primary and auxiliary healthcare facilities in developed countries help in maintaining healthy living standards past 65 years of age. Consequently, a cut-off of 65 years of age is used to define and study falls (and other health issues) in 'the elderly'. In contrast, the lack of proper healthcare facilities and unfavourable socio-economic conditions can drive quicker aging in developing and underdeveloped countries. Indeed, telomere length, a proxy for biological ageing, has been shown to be lower in harsh socio-economic conditions (Diez Roux et al. 2009) (Rewak et al. 2014) (Epel et al. 2004). These data propelled us to make a case for studying fall-risk in an Indian population that is biologically younger and yet more prone to falls.

As shown in supplementary figure 1, unintentional fall related injuries ranged from 1-9% in various age groups in India. The above 80 age group displayed the highest rate of falls at 8.84%. Whereas, less than 20-year olds displayed the lowest rate of falls at 1.13%. Despite a lack of significant differences between age groups, the rate of fall related-injury steadily rose after the age of 50. The rate of injury was 4.3% in the 50-60 age group, 4.7% in the 60-70 age group and

6.5% in the 70-80 age group. We observed a jump in the fall injuries in 3 specific age groups: the 20-30 age group (p=0.08), the 50-60 age group (p=0.15), the 70-80 age group (p=0.07). Given that the average Indian life expectancy at the time of this survey was 65 years of age (World Bank 2017), we decided to study fall related injuries in the aging Indian population at and after the second "jump in fall injuries" mark (50-60 years).

In the past, the WHO's SAGE team has studied ageing by gathering data from people aged 50+ in six LMIC (Low-Middle income countries). A 50 years of age cut-off has also been used in studying fall-risks in other developing countries (Stewart Williams et al. 2015).

*<u>Performance of the prediction models to predict fall-risk individuals aged 50+</u>*

Supplementary Table II: Performance values of the 4 fall-prediction models on the test data set.

| **Model Number** | Cut-off score in the development set | Specificity (%) | Sensitivity (%) | NPV (%) | PPV (%) |
|---|---|---|---|---|---|
| 1 | 12.5/34 | 63.98 | 85.48 | 98.83 | 11.00 |
| 2 | 0.0413/8.425 | 72.04 | 80.65 | 98.62 | 13.05 |
| 3 | 17.24/46.93 | 92.53 | 41.94 | 98.84 | 22.61 |
| 4 | 0.0100/3.32 | 67.93 | 82.26 | 98.65 | 11.78 |

Supplementary Table III: Performance values of the 4 fall-prediction models on training data and internal validation.

| **Model Number** | Cut-off score | AUC | AUC 95%CI with 1000x bootstrapping | Specificity (%) | Sensitivity (%) | NPV (%) | PPV (%) |
|---|---|---|---|---|---|---|---|
| 1 | 12.5/34 | 0.779 | 0.7494-0.8045 | 64.44 | 76.72 | 98.28 | 9.48 |
| 2 | 0.0413/8.425 | 0.790 | 0.7774-0.8281 | 68.17 | 75.86 | 98.31 | 10.37 |
| 3 | 17.24/46.93 | 0.790 | 0.7706-0.8255 | 67.41 | 77.16 | 98.38 | 10.31 |
| 4 | 0.0100/3.32 | 0.788 | 0.7749-0.8262 | 67.27 | 76.29 | 98.32 | 10.17 |

*Relative importance of variables present in the Indian Census data*

**Supplementary Table IV: Relative Importance of 6 variables determined in our data set that were available in the India Census 2011.**

| Variables | Relative Importance normalized to sum 100% |
|---|---|
| Hearing Problem | 22.18 |
| Married or not | 1.85 |
| Education - Primary school or less | 4.82 |
| Vision Problem | 15.55 |
| Gender | 26.37 |
| Not worked in previous 12 months | 29.23 |

Of the 6 variables common to the WHO data and the concensus data, relative importance of not having worked in the last 12 months was the highest (29.23%). This was followed by gender (26.37%), hearing (22.18%), vision (15.55%) and education (4.82%) and marital status (1.85%). These values were combined to give a "Risk val" using the procedure described above (see methods).
